# Hydroxychloroquine and azithromycin as a treatment of COVID-19: results of an open-label non-randomized clinical trial

**DOI:** 10.1101/2020.03.16.20037135

**Authors:** Philippe Gautret, Jean-Christophe Lagier, Philippe Parola, Van Thuan Hoang, Line Meddeb, Morgane Mailhe, Barbara Doudier, Johan Courjon, Valérie Giordanengo, Vera Esteves Vieira, Hervé Tissot Dupont, Stéphane Honoré, Philippe Colson, Eric Chabrière, Bernard La Scola, Jean-Marc Rolain, Philippe Brouqui, Didier Raoult

## Abstract

**Background:** Chloroquine and hydroxychloroquine have been found to be efficient on SARS-CoV-2, and reported to be efficient in Chinese COV-19 patients. We evaluate the role of hydroxychloroquine on respiratory viral loads.

**Patients and methods:** Patients were included in a single arm protocol to receive 600mg of hydroxychloroquine daily and their viral load in nasal swabs was tested daily. Depending on their clinical presentation, azithromycin was added to the treatment. Untreated patients from another center and cases refusing the protocol were included as negative controls. Presence and absence of virus at Day6-post inclusion was considered the end point.

**Results:** Twenty cases were treated in this study and showed a significant reduction of the viral carriage at D6-post inclusion compared to controls, and much lower average carrying duration than reported of untreated patients in the literature. Azithromycin added to hydroxychloroquine was significantly more efficient for virus elimination.

**Conclusion:** Hydroxychloroquine is significantly associated with viral load reduction/disappearance in COVID-19 patients and its effect is reinforced by azithromycin.

## 1. Introduction

In late December 2019, an outbreak of an emerging disease (COVID-19) due to a novel coronavirus (named SARS-CoV-2 latter) started in Wuhan, China and rapidly spread in China and outside [1,2]. The WHO declared the epidemic of COVID-19 as a pandemic on March 12^th^ 2020 [3]. According to a recent Chinese stud, about 80% of patients present with mild disease and the overall case-fatality rate is about 2.3% but reaches 8.0% in patients aged 70 to 79 years and 14.8% in those aged ≥80 years [4]. However, there is probably an important number of asymptomatic carriers in the population, and thus the mortality rate is probably overestimated. France is now facing the COVID-19 wave with more than 4500 cases, as of March 14^th^ 2020 [5]. Thus, there is an urgent need for an effective treatment to treat symptomatic patients but also to decrease the duration of virus carriage in order to limit the transmission in the community. Among candidate drugs to treat COVID-19, repositioning of old drugs for use as antiviral treatment is an interesting strategy because knowledge on safety profile, side effects, posology and drug interactions are well known [6,7].

A recent paper reported an inhibitor effect of remdesivir (a new antiviral drug) and chloroquine (an old antimalarial drug) on the growth of SARS-CoV-2 *in vitro*, [8] and an early clinical trial conducted in COVID-19 Chinese patients, showed that chloroquine had a significant effect, both in terms of clinical outcome and viral clearance, when comparing to controls groups [9,10]. Chinese experts recommend that patients diagnosed as mild, moderate and severe cases of COVID-19 pneumonia and without contraindications to chloroquine, be treated with 500 mg chloroquine twice a day for ten days [11].

Hydroxychloroquine (an analogue of chloroquine) has been demonstrated to have an anti-SARS-CoV activity *in vitro* [12]. Hydroxychloroquine clinical safety profile is better than that of chloroquine (during long-term use) and allows higher daily dose [13] and has fewer concerns about drug-drug interactions [14]. Our team has a very comprehensive experience in successfully treating patients with chronic diseases due to intracellular bacteria (Q fever due to *Coxiella burnetii* and Whipple’s disease due to *Tropheryma whipplei*) with long-term hydroxychloroquine treatment (600 mg/day for 12 to 18 months) since more than 20 years. [15,16] We therefore started to conduct a clinical trial aiming at assessing the effect of hydroxychloroquine on SARS-CoV-2-infected patients after approval by the French Ministry of Health. In this report we describe our early results, focusing on virological data in patients receiving hydroxychloroquine as compared to a control group.

## 2. Study population and Methods

### Setting

This ongoing study is coordinated by The Méditerranée Infection University Hospital Institute in Marseille. Patients who were proposed a treatment with hydroxychloroquine were recruited and managed in Marseille centre. Controls without hydroxychloroquine treatment were recruited in Marseille, Nice, Avignon and Briançon centers, all located in South France.

### Patients

Hospitalized patients with confirmed COVID-19 were included in this study if they fulfilled two primary criteria: i) age >12 years; ii) PCR documented SARS-CoV-2 carriage in nasopharyngeal sample at admission whatever their clinical status.

Patients were excluded if they had a known allergy to hydroxychloroquine or chloroquine or had another known contraindication to treatment with the study drug, including retinopathy, G6PD deficiency and QT prolongation. Breastfeeding and pregnant patients were excluded based on their declaration and pregnancy test results when required.

### Informed consent

Before being included in the study, patients meeting inclusion criteria had to give their consent to participate to the study. Written informed signed consent was obtained from adult participants (≥ 18 years) or from parents or legal guardians for minors (<18 years). An information document that clearly indicates the risks and the benefits associated with the participation to the study was given to each patient. Patients received information about their clinical status during care regardless of whether they participate in the study or not. Regarding patient identification, a study number was assigned sequentially to included participants, according to the range of patient numbers allocated to each study centre. The study was conducted in accordance with the International Council for Harmonisation of Technical Requirements for Pharmaceuticals for Human Use (ICH) guidelines of good clinical practice, the Helsinki Declaration, and applicable standard operating procedures.

The protocol, appendices and any other relevant documentation were submitted to the French National Agency for Drug Safety (ANSM) (2020-000890-25) and to the French Ethic Committee (CPP Ile de France) (20.02.28.99113) for reviewing and approved on 5^th^ and 6^th^ March, 2020, respectively. This trial is registered with EU Clinical Trials Register, number 2020-000890-25.

### Procedure

Patients were seen at baseline for enrolment, initial data collection and treatment at day-0, and again for daily follow-up during 14 days. Each day, patients received a standardized clinical examination and when possible, a nasopharyngeal sample was collected. All clinical data were collected using standardized questionnaires. All patients in Marseille center were proposed oral hydroxychloroquine sulfate 200 mg, three times per day during ten days (in this preliminary phase, we did not enrolled children in the treatment group based in data indicating that children develop mild symptoms of COVID-19 [4]). Patients who refused the treatment or had an exclusion criteria, served as controls in Marseille centre. Patients in other centers did not receive hydroxychloroquine and served as controls. Symptomatic treatment and antibiotics as a measure to prevent bacterial super-infection was provided by investigators based on clinical judgment. Hydroxychloroquine was provided by the National Pharmacy of France on nominative demand.

### Clinical classification

Patients were grouped into three categories: asymptomatic, upper respiratory tract infection (URTI) when presenting with rhinitis, pharyngitis, or isolated low-grade fever and myalgia, and lower respiratory tract infections (LRTI) when presenting with symptoms of pneumonia or bronchitis.

### PCR assay

SARS-CoV-2 RNA was assessed by real-time reverse transcription-PCR [17].

### Hydroxychloroquine dosage

Native hydroxychloroquine has been dosed from patients’ serum samples by UHPLC-UV using a previously described protocol [18]. The peak of the chromatogram at 1.05 min of retention corresponds to hydroxychloroquine metabolite. The serum concentration of this metabolite is deduced from UV absorption, as for hydroxychloroquine concentration. Considering both concentrations provides an estimation of initial serum hydroxychloroquine concentration.

### Culture

For all patients, 500 µL of the liquid collected from the nasopharyngeal swab were passed through 0.22-µm pore sized centrifugal filter (Merck millipore, Darmstadt, Germany), then were inoculated in wells of 96-well culture microplates, of which 4 wells contained Vero E6 cells (ATCC CRL-1586) in Minimum Essential Medium culture medium with 4% fetal calf serum and 1% glutamine. After centrifigation at 4,000 g, microplates were incubated at 37°C. Plates were observed daily for evidence of cytopathogenic effect. Presumptive detection of virus in supernatant was done using SU5000 SEM (Hitachi) then confirmed by specific RT-PCR.

### Outcome

The primary endpoint was virological clearance at day-6 post-inclusion. Secondary outcomes were virological clearance overtime during the study period, clinical follow-up (body temperature, respiratory rate, long of stay at hospital and mortality), and occurrence of side-effects.

### Statistics

Assuming a 50% efficacy of hydroxychloroquine in reducing the viral load at day 7, a 85% power, a type I error rate of 5% and 10% loss to follow-up, we calculated that a total of 48 COVID-19 patients (ie, 24 cases in the hydroxychloroquine group and 24 in the control group) would be required for the analysis (Fleiss with CC). Statistical differences were evaluated by Pearson’s chi-square or Fisher’s exact tests as categorical variables, as appropriate. Means of quantitative data were compared using Student’s t-test. Analyses were performed in OpenEpi version 3.0.

## 3. Results (detailed results are available in supplementary Table 1)

### Demographics and clinical presentation

We enrolled 36 out of 42 patients meeting the inclusion criteria in this study that had at least six days of follow-up at the time of the present analysis. A total of 26 patients received hydroxychloroquine and 16 were control patients. Six hydroxychloroquine-treated patients were lost in follow-up during the survey because of early cessation of treatment. Reasons are as follows: three patients were transferred to intensive care unit, including one transferred on day2 post-inclusion who was PCR-positive on day1, one transferred on day3 post-inclusion who was PCR-positive on days1-2 and one transferred on day4 post-inclusion who was PCR-positive on day1 and day3; one patient died on day3 post inclusion and was PCR-negative on day2; one patient decided to leave the hospital on day3 post-inclusion and was PCR-negative on days1-2; finally, one patient stopped the treatment on day3 post-inclusion because of nausea and was PCR-positive on days1-2-3. The results presented here are therefore those of 36 patients (20 hydroxychloroquine-treated patients and 16 control patients). None of the control patients was lost in follow-up. Basic demographics and clinical status are presented in Table 1. Overall, 15 patients were male (41.7%), with a mean age of 45.1 years. The proportion of asymptomatic patients was 16.7%, that of patients with URTI symptoms was 61.1% and that of patients with LRTI symptoms was 22.2%). Hydroxychloroquine-treated patients were older than control patients (51.2 years vs. 37.3 years). No significant difference was observed between hydroxychloroquine-treated patients and control patients with regard to gender, clinical status and duration of symptoms prior to inclusion (Table 1). Among hydroxychloroquine-treated patients six patients received azithromycin to prevent bacterial super-infection under daily electrocardiogram control. Clinical follow-up and occurrence of side-effects will be described in a further paper at the end of the trial.

**Table 1.** Characteristics of the study population.

|  | Age (years) |  |  | Male gender |  | Clinical status |  |  |  | Time between onset of symptoms and inclusion (days) |  |  |
| --- | --- | --- | --- | --- | --- | --- | --- | --- | --- | --- | --- | --- |
| | Mean ± SD | t | p-value | n (%) | p-value | Asymptomatic | URTI | LRTI | p-value | Mean ± SD | t | p-value |
| Hydroxychloroquine treated patients (N=20) | 51.2 ± 18.7 | -1.95 | 0.06 | 9 (45.0) | 0.65 | 2 (10.0) | 12 (60.0) | 6 (30.0) | 0.30 | 4.1 ± 2.6 | -0.15 | 0.88 |
| Control patients (N=16) | 37.3 ± 24.0 | | | 6 (37.5) | | 4 (25.0) | 10 (62.5) | 2 (12.5) | | 3.9 ± 2.8 | | |
| All patients (36) | 45.1 ± 22.0 | | | 15 (41.7) | | 6 (16.7) | 22 (61.1) | 8 (22.2) | | 4.0 ± 2.6 | | |

URTI: upper tract respiratory infection, LRTI: lower tract respiratory infection

### Hydroxychloroquine dosage

Mean hydroxychloroquine serum concentration was 0.46 µg/ml±0.2 (N=20).

### Effect of hydroxychloroquine on viral load

The proportion of patients that had negative PCR results in nasopharyngeal samples significantly differed between treated patients and controls at days 3-4-5 and 6 post-inclusion (Table 2). At day6 post-inclusion, 70% of hydroxychloroquine-treated patients were virologicaly cured comparing with 12.5% in the control group (p= 0.001).

**Table 2.** Proportion of patients with virological cure (negative nasopharyngeal PCR) by day, in COVID-19 patients treated with hydroxychloroquine and in COVID-19 control patients.

|  | Day3 post inclusion |  |  | Day4 post inclusion |  |  | Day5 post inclusion |  |  | Day6 post inclusion |  |  |
| --- | --- | --- | --- | --- | --- | --- | --- | --- | --- | --- | --- | --- |
|  | Number of<br>negative<br>patients/total<br>number of<br>patients | % | p-value | Number of<br>negative<br>patients/total<br>number of<br>patients | % | p-value | Number of<br>negative<br>patients/total<br>number of<br>patients | % | p-value | Number of<br>negative<br>patients/total<br>number of<br>patients | % | p-value |
| Hydroxychloroquine<br>treated patients<br>(N=20) | 10/20 | 50.0 | 0.005 | 12/20 | 60.0 | 0.04 | 13/20 | 65.0 | 0.006 | 14/20 | 70.0 | 0.001 |
| Control patients<br>(N=16) | 1/16 | 6.3 |  | 4/16 | 25.0 |  | 3/16 | 18.8 |  | 2/16 | 12.5 |  |

<sup>a</sup>control patients from centers other than Marseille did not underwent daily sampling, but were sampled every other day, they were considered positive for PCR when actually positive the day before an the day after the day with missing data.

**Table 3.** Proportion of patients with virological cure (negative nasopharyngeal PCR) by day, in COVID-19 patients treated with hydroxychloroquine only, in COVID-19 patients treated with hydroxychloroquine and azithomycin combination, and in COVID-19 control patients.

|  | Day3 post inclusion |  |  | Day4 post inclusion |  |  | Day5 post inclusion |  |  | Day6 post inclusion |  |  |
| --- | --- | --- | --- | --- | --- | --- | --- | --- | --- | --- | --- | --- |
|  | Number of negative patients/total number of patients | % | p-value | Number of negative patients/total number of patients | % | p-value | Number of negative patients/total number of patients | % | p-value | Number of negative patients/total number of patients | % | p-value |
| Control patients | 1/16 | 6.3 | 0.002 | 4/16 | 25.0 | 0.05 | 3/16 | 18.8 | 0.002 | 2/16 | 12.5 | <0.001 |
| Hydroxychloroquine treatment only | 5/14 | 35.7 |  | 7/14 | 50.0 |  | 7/14 | 50.0 |  | 8/14 | 57.1 |  |
| Hydroxychloroquine and azithromycin combined treatment | 5/6 | 83.3 |  | 5/6 | 83.3 |  | 6/6 | 100 |  | 6/6 | 100 |  |

When comparing the effect of hydroxychloroquine treatment as a single drug and the effect of hydroxychloroquine and azithromyc in combination, the proportion of patients that had negative PCR results in nasopharyngeal samples was significantly different between the two groups at days 3-4-5 and 6 post-inclusion. At day6 post-inclusion, 100% of patients treated with hydroxychloroquine and azithromycin combination were virologicaly cured comparing with 57.1% in patients treated with hydroxychloroquine only, and 12.5% in the control group (p<0.001). These results are summarized in Figures 1 and 2.

**Figure 1.**
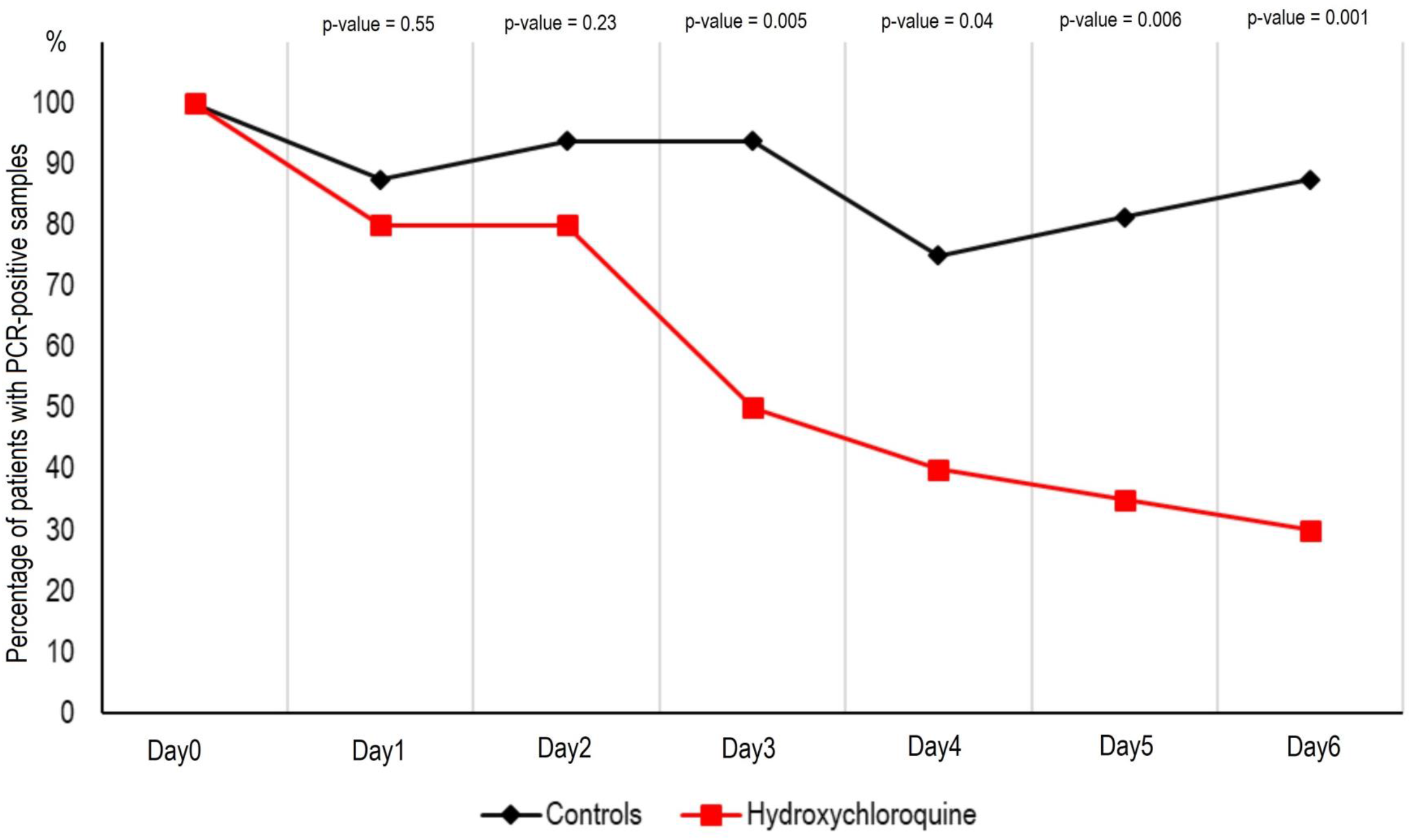
Percentage of patients with PCR-positive nasopharyngeal samples from inclusion to day6 post-inclusion in COVID-19 patients treated with hydroxychloroquine and in COVID-19 control patients.

**Figure 2.**
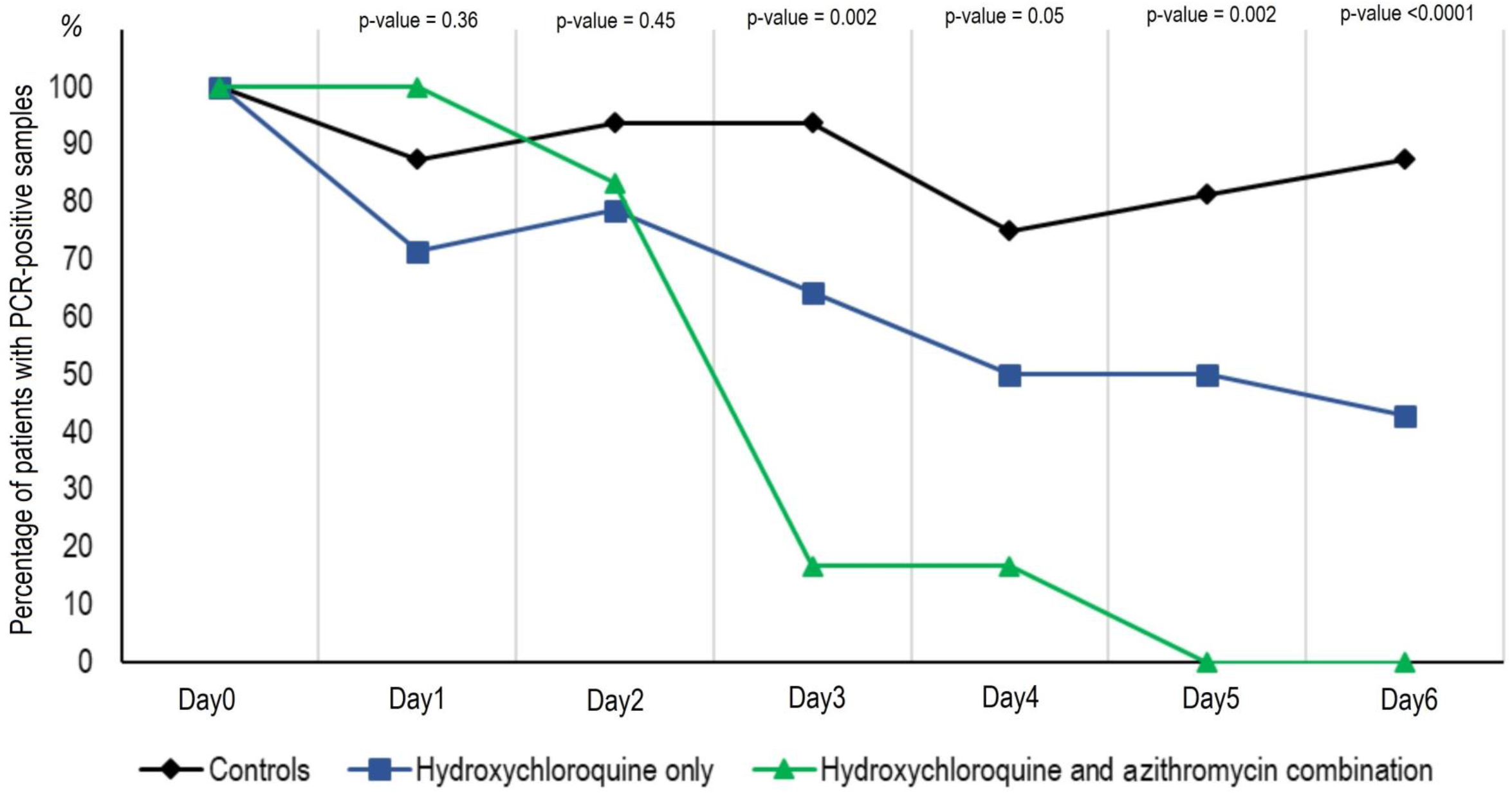
Percentage of patients with PCR-positive nasopharyngeal samples from inclusion to day6 post-inclusion in COVID-19 patients treated with hydroxychloroquine only, in COVID-19 patients treated with hydroxychloroquine and azithomycin combination, and in COVID-19 control patients.

Of note, one patient who was still PCR-positive at day6-post inclusion under hydroxychloroquine treatment only, received azithromycin in addition to hydroxychloroquine at day8-post inclusion and cured her infection at day-9 post infection. In contrast, one of the patients under hydroxychloroquine and azithromycin combination who tested negative at day6 post-inclusion was tested positive at low titer at day8 post-inclusion.

### Cultures

We could isolate SARS-CoV-2 in 19 out of 25 clinical samples from patients.

## 4. Discussion

For ethical reasons and because our first results are so significant and evident we decide to share our findings with the medical community, given the urgent need for an effective drug against SARS-CoV-2 in the current pandemic context.

We show here that hydroxychloroquine is efficient in clearing viral nasopharyngeal carriage of SARS-CoV-2 in COVID-19 patients in only three to six days, in most patients. A significant difference was observed between hydroxychloroquine-treated patients and controls starting even on day3 post-inclusion. These results are of great importance because a recent paper has shown that the mean duration of viral shedding in patients suffering from COVID-19 in China was 20 days (even 37 days for the longest duration) [19]

Very recently, a Chinese team published results of a study demonstrating that chloroquine and hydroxychloroquine inhibit SARS-CoV-2 *in vitro* with hydroxychloroquine (EC50=0.72%µM) found to be more potent than chloroquine (EC50=5.47%µM) [14]. These *in vitro* results corroborate our clinical results. The target values indicated in this paper [14] were reached in our experiments. The safer dose-dependent toxicity profile of hydroxychloroquine in humans, compared to that of chloroquine [13] allows using clinical doses of hydroxychloroquine that will be over its EC50 observed *in vitro* [14].

Our preliminary results also suggest a synergistic effect of the combination of hydroxychloroquine and azithromycin. Azithromycin has been shown to be active *in vitro* against Zika and Ebola viruses [20–22] and to prevent severe respiratory tract infections when administrated to patients suffering viral infection [23]. This finding should be further explored to know whether a combination is more effective especially in severe cases. Speculated potential risk of severe QT prolongation induced by the association of the two drugs has not been established yet but should be considered. As for each treatment, the cost benefits of the risk should be evaluated individually. Further studies on this combination are needed, since such combination may both act as an antiviral therapy against SARS-CoV-2 and prevent bacterial super-infections.

The cause of failure for hydroxychloroquine treatment should be investigated by testing the isolated SARS-CoV-2 strains of the non-respondents and analyzing their genome, and by analyzing the host factors that may be associated with the metabolism of hydroxychloroquine. The existence of hydroxychloroquine failure in two patients (mother and son) is more suggestive of the last mechanism of resistance.

Such results are promising and open the possibility of an international strategy to decision-makers to fight this emerging viral infection in real-time even if other strategies and research including vaccine development could be also effective, but only in the future. We therefore recommend that COVID-19 patients be treated with hydroxychloroquine and azithromycin to cure their infection and to limit the transmission of the virus to other people in order to curb the spread of COVID-19 in the world. Further works are also warranted to determine if these compounds could be useful as chemoprophylaxis to prevent the transmission of the virus, especially for healthcare workers.

## Data Availability

All data are available on request

## Acknowledgements

We thank Céline Boschi, Stéphanie Branger, Véronique Filosa, Géraldine Gonfier, Nadège Palmero, Magali Richez and all the clinical, technical and paramedical staffs of the hospitalization units and laboratories for their support in this difficult context.

## Funding source

This work was supported by the French Government under the « Investissements d’avenir » (Investments for the Future) program managed by the Agence Nationale de la Recherche (ANR, fr: National Agency for Research), (reference: Méditerranée Infection 10-IAHU-03)

---

**Supplementary Table 1.**

| Patient | Age (years) | Sex | Clinical status | Time between onset of symptoms and inclusion (days) | Hydroxychloroquine treatment | Hydroxychloroquine serum concentration µg/ml (day of dosage) | Azithromycin treatment | D0 | D1 | D2 | D3 | D4 | D5 | D6 |
| --- | --- | --- | --- | --- | --- | --- | --- | --- | --- | --- | --- | --- | --- | --- |
|  | 10 | M | Asymptomatic | - | No | - | No | 31 | NEG | NEG | NEG | NEG | NEG | NEG |
|  | 12 | F | Asymptomatic | - | No | - | No | 26 | ND | 33 | 34 | NEG | 34 | NEG |
|  | 14 | F | Asymptomatic | - | No | - | No | 26 | 31 | 23 | 22 | 27 | NEG | 26 |
|  | 10 | M | Asymptomatic | - | No | - | No | 24 | NEG | 33 | 33 | NEG | NEG | 32 |
|  | 20 | M | URTI | 4 | No | - | No | 24 | 24 | 24 | 27 | NEG | 31 | 29 |
|  | 65 | F | URTI | 2 | No | - | No | POS | ND | POS | ND | POS | ND | POS |
|  | 46 | M | URTI | Unknown | No | - | No | 28 | NF | NF | NF | 26 | NF | 30 |
|  | 69 | M | LRTI | 2 | No | - | No | POS | POS | POS | POS | POS | POS | POS |
|  | 62 | F | LRTI | 10 | No | - | No | POS | ND | POS | ND | POS | ND | POS |
|  | 66 | F | URTI | 0 | No | - | No | POS | ND | POS | ND | POS | ND | POS |
|  | 75 | F | URTI | 3 | No | - | No | POS | ND | POS | ND | POS | ND | POS |
|  | 23 | F | URTI | 5 | No | - | No | POS | ND | POS | ND | POS | ND | POS |
|  | 45 | F | URTI | Unknown | No | - | No | POS | ND | POS | ND | POS | ND | POS |
|  | 16 | M | URTI | 2 | No | - | No | POS | ND | ND | POS | ND | POS | POS |
|  | 42 | F | URTI | 5 | No | - | No | ND | POS | ND | POS | ND | POS | POS |
|  | 23 | F | URTI | 6 | No | - | No | ND | POS | ND | POS | ND | POS | POS |
|  | 44 | F | URTI | 6 | Yes | 0.519 (D6) | No | 30 | ND | 29 | 26 | 32 | 26 | 31 |
|  | 54 | M | Asymptomatic | - | Yes | 0.462 (D6) | No | 29 | NEG | NEG | NEG | NEG | NEG | NEG |
|  | 25 | M | URTI | 3 | Yes | 0.419 (D6) | No | 23 | 25 | 28 | 25 | NEG | NEG | NEG |
|  | 59 | F | Asymptomatic | - | Yes | 0.288 (D4) | No | 30 | NEG | NEG | NEG | NEG | NEG | NEG |
|  | 49 | F | URTI | 1 | Yes | 0.621 (D6) | No | 34 | 27 | 19 | 16 | 34 | 24 | 22 |
|  | 24 | F | URTI | 10 | Yes | 0.723 (D6) | No | 28 | NEG | 32 | 34 | NEG | NEG | NEG |
|  | 81 | F | LRTI | 2 | Yes | 0.591 (D6) | No | 22 | 21 | 30 | NEG | 32 | 28 | NEG |
|  | 85 | F | LRTI | 1 | Yes | 0.619 (D6) | No | 17 | 21 | 23 | 21 | 26 | 24 | 24 |
|  | 40 | M | URTI | 3 | Yes | 0.418 (D6) | No | 22 | NF | 28 | 21 | 15 | 20 | 17 |
|  | 53 | M | URTI | 5 | Yes | 0.515 (D6) | No | 27 | 28 | 32 | 31 | NEG | NEG | NEG |
|  | 63 | F | URTI | 8 | Yes | 0.319 (D4) | No | 34 | NEG | 30 | NEG | NEG | NEG | NEG |
|  | 42 | F | URTI | 1 | Yes | 0.453 (D6) | No | 19 | 16 | 17 | 17 | 19 | 20 | 31 |
|  | 87 | F | URTI | 5 | Yes | 0.557 (D6) | No | 25 | 30 | NEG | NEG | NEG | ND | ND |
|  | 33 | M | URTI | 2 | Yes | 0.194 (D2) | No | 15 | 23 | 26 | 26 | NF | 32 | 32 |
|  | 53 | F | LRTI | 7 | Yes | 1.076 (D6) | Yes | 28 | 31 | 34 | NEG | 34 | NEG | NEG |
|  | 48 | M | URTI | 2 | Yes | 0.57 (D6) | Yes | 23 | 29 | 29 | NEG | NEG | NEG | NEG |
|  | 50 | F | LRTI | 5 | Yes | 0.827 (D6) | Yes | 30 | 27 | NEG | NEG | NEG | NEG | NEG |
|  | 20 | M | URTI | 2 | Yes | 0.381 (D6) | Yes | 27 | 31 | 29 | NEG | NEG | NEG | NEG |
|  | 54 | M | LRTI | 6 | Yes | 0.366 (D4) | Yes | 24 | ND | ND | 29 | NEG | NEG | NEG |
|  | 60 | M | LRTI | 4 | Yes | 0.319 (D4) | Yes | 29 | 31 | 31 | NEG | NEG | NEG | NEG |

URTI: upper tract respiratory infection, LRTI: lower tract respiratory infection, POS: positive PCR, NEG, negative PCR, ND, PCR not done

## Notes

### Competing Interest Statement

The authors have declared no competing interest.

### Clinical Trial

This clinical trial was approved by the French National Agency for Drug safety (ANSM) (2020-000890-25) and to the French Ethic Committee (CPP Ile de France) (20.02.28.99113) for reviewing and approved on 5th and 6th March, 2020, respectively. This trial is registered with EU Clinical trials register number 2020-000890-25.

